# Health Impact Assessment of BRCA1/2 Cascade Screening for the Personalized Prevention of Hereditary Breast and Ovarian Cancers in Italy

**DOI:** 10.64898/2026.04.13.26350758

**Authors:** Angelica Valz Gris, Erika Giacobini, Vittoria Tricomi, Filippo Rumi, Ilaria Valentini, Antonio Cristiano, Salvatore Testa, Aldo Rosano, Angelo Maria Pezzullo, Stefania Boccia

## Abstract

**Introduction:** Pathogenic germline variants in the *BRCA1* and *BRCA2* genes confer a markedly increased risk of breast and ovarian cancer, for which effective preventive strategies are available. Although national and international guidelines recommend *BRCA* testing and cascade screening of relatives, implementation in Italy remains highly heterogeneous across regions. This study estimates the potential population health and cost impact of achieving full nationwide implementation of *BRCA1/2* cascade screening in Italy and identifies key organisational barriers and priority actions for implementation.

**Methods:** We conducted a Health Impact Assessment integrating literature review, simulation modelling, and stakeholder consultation. A decision tree and Markov model compared the current heterogeneous implementation of *BRCA* screening in Italy with an ideal scenario reflecting full adherence to national guidelines, optimal cascade screening, and uptake of preventive strategies. Outcomes included breast and ovarian cancer incidence and mortality, healthcare costs over a lifetime horizon (80 years). Key barriers affecting organisational feasibility, acceptability, and patient well-being were assessed, and a set of priority action recommendations was developed.

**Results:** In the ideal scenario, 25,626 eligible cancer patients would undergo *BRCA* testing annually, identifying 4,254 mutation carriers and enabling cascade testing of 27,650 relatives, of whom 8,682 would be *BRCA*-positive. Under the current implementation, only 8,807 patients and 2,168 relatives are tested, identifying 948 carriers. Over 30 years, full implementation would prevent 821 cancer cases (–27.9%) and 1,282 deaths (–49.7%) compared with the current scenario. While initial expenditures increase due to expanded testing and preventive interventions, cumulative costs decrease over time, resulting in net savings of €5.8 million at 30 years and a saving per event avoided (–€2,779). Major implementation barriers include fragmented governance, limited access to genetic counselling, heterogeneous laboratory practices, insufficient professional training, and weak referral pathways.

**Conclusion:** Full implementation of *BRCA1/2* cascade screening in Italy would yield substantial population health benefits and long-term cost savings. Coordinated national governance, standardized pathways, investment in counselling and workforce capacity, and robust monitoring systems are essential to ensure equitable access and sustainable delivery of personalized cancer prevention. This study demonstrates the value of the HIA methodology for evaluating and guiding genomic prevention policies.

## Introduction

Pathogenic germline variants in the *BRCA1* and *BRCA2* genes are among the most well-characterised genetic contributors to cancer susceptibility [1]. These mutations significantly impair the cellular capacity to repair DNA damage via homologous recombination, resulting in genomic instability and a markedly increased risk of developing several malignancies. In women, *BRCA1/2* mutations are responsible for approximately 5–10% of all breast cancers and 15–20% of epithelial ovarian cancers, with cumulative lifetime risks reaching up to 70% for breast cancer and 60% for ovarian cancer, depending on the gene involved and other factors [2]. Additional associations have been documented with pancreatic, prostate, and other less frequent cancer types.

Although *BRCA1/2*-associated cancers account for a limited proportion of overall breast and ovarian cancer cases, their clinical relevance is underscored by the high penetrance of the underlying mutations and the availability of evidence-based risk-reduction strategies. The identification of unaffected carriers through predictive testing enables the adoption of preventive interventions with demonstrated efficacy [3]. These include surveillance protocols, aimed at early detection, and prophylactic surgical procedures, such as risk-reducing salpingo-oophorectomy (RRSO) and risk-reducing mastectomy (RRM), which have been associated with significant reductions in cancer incidence and disease-specific mortality in high-risk populations [4].

International scientific and professional societies have long recognised the importance of *BRCA1/2* testing in hereditary cancer prevention and developed clinical guidelines that are continuously updated to reflect the latest evidence on the effectiveness of genetic testing and preventive interventions [5–8]. *BRCA* testing was first introduced in the 2007 guidelines of the Italian Association of Medical Oncology for breast and ovarian cancer [9]. The 2014–2018 Italian National Prevention Plan subsequently established the first national framework for *BRCA1/2* testing in hereditary breast and ovarian cancer and required regions to develop structured genetic testing programs [10]. Despite these initiatives, implementation of *BRCA1/2* testing and management remains heterogeneous across regions. Many regions have developed Diagnostic, Therapeutic, and Assistance Pathways (PDTAs) to operationalise national recommendations, but these differ markedly in structure, scope, and integration with cancer care. Key implementation components, including reimbursement for testing and preventive care, legal and regulatory provisions, workforce training, and public awareness, remain insufficient in several settings, reflecting broader implementation challenges. In

October 2025, the State–Regions Conference reached an initial agreement to update the Italian Essential Levels of Care, the set of services guaranteed by the Italian National Health Service, formally including *BRCA* genetic testing and surveillance pathways for hereditary breast and ovarian cancer [11]. Although the approval of this agreement would represent a key step toward more comprehensive implementation, several barriers still hinder the full rollout of nationwide cascade screening. This study evaluates the potential clinical and societal impact of nationwide *BRCA1/2* cascade screening in Italy by estimating population-level effects on breast and ovarian cancer incidence and mortality and identifying key organisational barriers to equitable implementation.

## Methods

We conducted a Health Impact Assessment (HIA) to evaluate the population health impact and system-level implications of standardising preventive *BRCA* testing in healthy high-risk individuals across Italian regions in line with national guidelines [12]. Testing in cancer patients for treatment planning was not considered within the scope of this HIA. A multidisciplinary Steering Committee (SC) was formed to guide the process. This SC included 1 medical geneticist, 1 oncologist, 1 radiologist, 1 general and reconstructive surgeon, 1 public health official from regional prevention departments, 1 laboratory professional, 1 representative of patient organisations, and 1 member of the private sector. Four thematic domains were defined to guide the assessment of policy impacts and implementation determinants. These included population health and well-being, economic sustainability, patient acceptability and awareness, and organisational feasibility. The proposed domains and associated assessment methodologies were discussed and validated by the SC.

### Evidence Assessment

The assessment used an integrated approach combining narrative literature reviews, simulation modelling, and stakeholder consultation through structured questionnaires and meetings. We searched for systematic reviews and clinical guidelines assessing the effectiveness of preventive strategies in individuals with *BRCA* pathogenic variants to support the development of the simulation model. We also retrieved systematic reviews on the economic impact of different testing strategies, as well as economic evaluations of *BRCA* cascade screening conducted in Italy. Moreover, we reviewed primary studies conducted in Italy that assessed patient acceptability and awareness and identified potential organisational, feasibility, and implementation barriers. Subsequently, we developed a simulation model to estimate the impact of implementing the proposed preventive strategies in Italy, in terms of the number of ovarian and breast cancer cases and related deaths avoided, as well as the associated costs. In parallel, consultations with the SC were conducted through meetings and a dedicated questionnaire, both to inform the development of the simulation model and to collect additional information on the current level of implementation, organisational barriers, and potential challenges. The evidence generated from the literature reviews, the simulation model, and the consultations was then synthesised and discussed with the SC.

### Simulation model

We compared two scenarios: 1) a current scenario reflecting the heterogeneous status of *BRCA* testing and preventive strategies implementation in Italy, and 2) an ideal scenario modelling a structured, standardised national program in which all eligible high-risk individuals are tested and receive the appropriate preventive strategies according to best practices.

To estimate the number of high-risk individuals tested under each scenario, we first estimated the annual number of cancer patients eligible for *BRCA* testing, from whom cascade screening would then extend to healthy relatives. In the ideal scenario, the eligible population comprised individuals with breast, ovarian, prostate, or pancreatic cancer who met the most recent national guideline criteria. We first estimated the proportion of cancer patients meeting each criterion using U.S. SEER data restricted to Caucasian individuals, selected for its individual-level detail needed to address overlap across criteria, which was not possible with Italian sources [13]; for family-history-based eligibility criteria in breast cancer, we used published estimates **(Supplementary Table 1)**. These proportions were applied to the Italian incidence for the four cancers from AIOM I numeri del Cancro 2023 [14]. For the current scenario, *BRCA* testing among high-risk patients was estimated from the inclusion of each eligibility criterion in regional PDTAs **(Supplementary Table 2)** and combined with regional incidence data to project the number of patients currently tested under each regional protocol. Following the estimation of the two high-risk populations, we developed a simulation model composed of two components. The first is a decision tree model describing testing and preventive strategies under the current and ideal implementation scenarios (**Figure 1**). In the current scenario, only the proportion of high-risk patients previously estimated based on the regional eligibility criteria is assumed to undergo *BRCA* testing. Among the *BRCA*-positive patients identified, some healthy family members may subsequently be tested, while others may not. Female carriers identified through cascade testing may follow different preventive pathways, including RRM, RRSO, active surveillance, or no personalised preventive intervention. Current uptake rates of *BRCA* testing and preventive interventions were derived from studies conducted in Italy and are reported in **Table 1** alongside all other parameters used in the model. In the ideal scenario, all high-risk patients and all eligible healthy relatives of *BRCA*-positive individuals are assumed to undergo testing, with all female *BRCA* carriers choosing one of the available preventive strategies, either surgical or surveillance-based.

**Figure 1.**
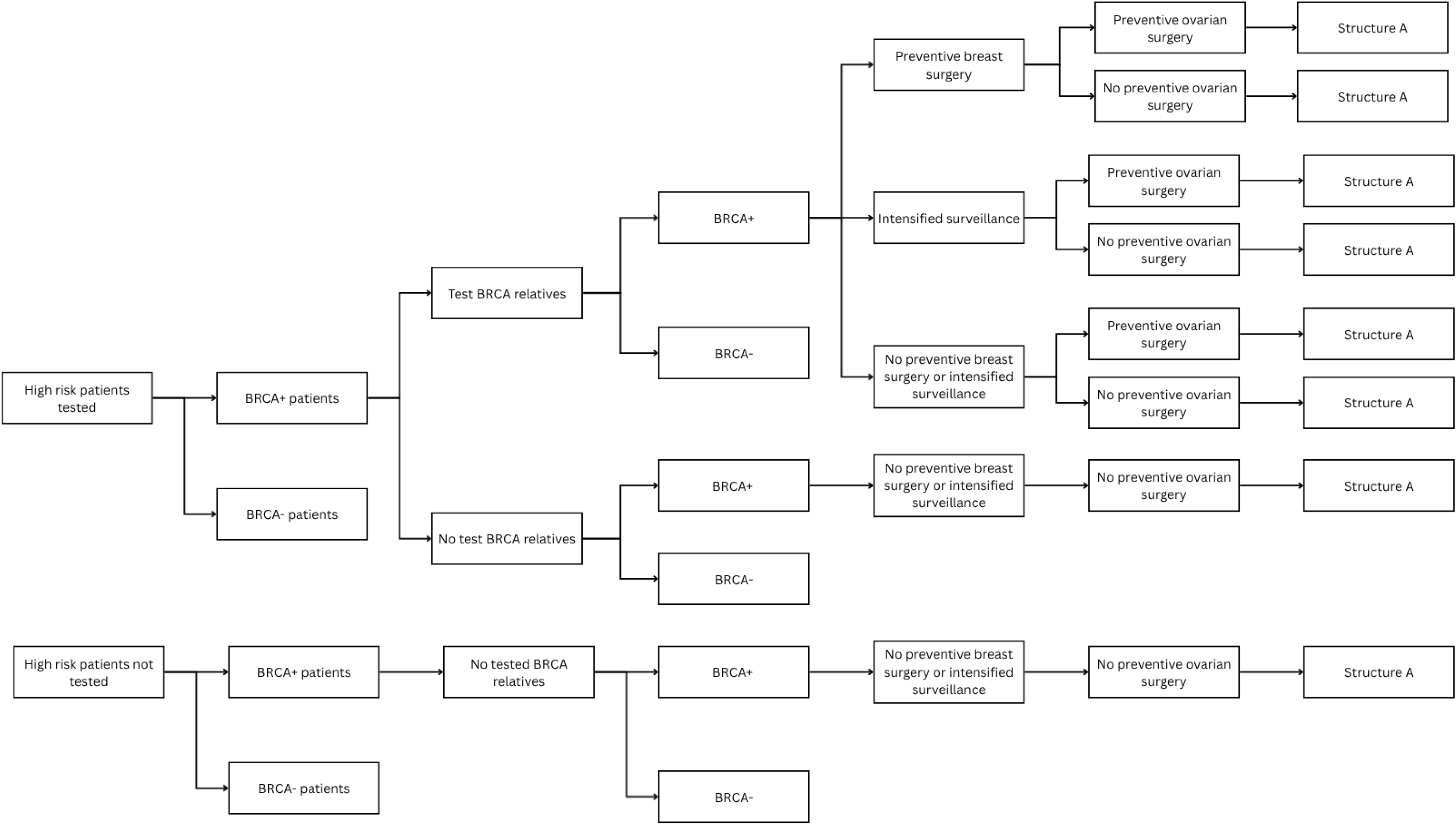
Simulation model. Decision tree model component.

**Table 1.** Data and sources used in the decision tree model.

| Description | Current Scenario | Ideal Scenario | Source |
| --- | --- | --- | --- |
| Percentage of carriers in high-risk patients | 16,6% |  | Alsop et al., 2012 [45] |
| Estimated number of first-degree relatives per positive patients | 2,66 |  | Trevisan et al., 2024 [37] |
| Estimated number of second-degree relatives per positive patients | 1,86 |  | Trevisan et al., 2024 [37] |
| Estimate number of third-degree relatives per positive patients | 1,97 |  | Trevisan et al., 2024 [37] |
| Percentage of First degree relative tested | 44,4% | 100% | Trevisan et al., 2024 [37] |
| Percentage of Second degree relative tested | 8,6% | 100% | Trevisan et al., 2024 [37] |
| Percentage of Third degree relative tested | 7,1% | 100% | Trevisan et al., 2024 [37] |
| Percentage of female carriers that undergo breast preventive surgery | 10% | 35,9% | Metcalfe et al, 2019 [40] |
| Percentage of female carriers that undergo active surveillance | 77,8% | 100% | Metcalfe et al, 2019 [40] |
| Percentage of female carriers that do not undergo to any personalized preventive intervention | 12% | 0% | Metcalfe et al, 2019 [40] |
| Percentage of female carriers that undergo ovarian preventive surgery | 52,5% | 55% | Metcalfe et al, 2019 [40] |
|  |  |  | Rebbeck et al, 2019 [4] |

All branches of the decision tree then converge into a common downstream Markov model (**Figure 2**), which is populated by the cohorts generated in the decision tree has been used to estimate the long-term effects of preventive strategies in *BRCA*-positive individuals in terms of cancer incidence, mortality, and costs. The model includes six initial management/prevention states: RRM and RRSO; RRM without RRSO; RRSO without RRM with active surveillance; no RRM and no RRSO with active surveillance; RRSO without RRM without active surveillance; and no RRM, no RRSO, and no active surveillance. From each of these states, individuals may remain in the same state in subsequent cycles, as indicated by the circular arrows, or transition to the next state. Cancer-related states include *BRCA*-associated breast cancer (*BRCA* BC) and *BRCA*-associated ovarian cancer (*BRCA* OC). Cancer-free states are defined as no breast cancer (no BC) and no ovarian cancer (no OC). From all states, including cancer states, transitions to the absorbing state of death are allowed, capturing both all-cause mortality and disease-specific mortality. The model aggregates probabilities and utilities assigned to each health state and generates mean expected values for costs and effects across the simulated population. To adequately reflect the long-term impact of hereditary cancer prevention, the model employed a one-year cycle length and an 80-year time horizon (lifetime). Data used in the Markov model are available in **Table 2**.

**Figure 2.**
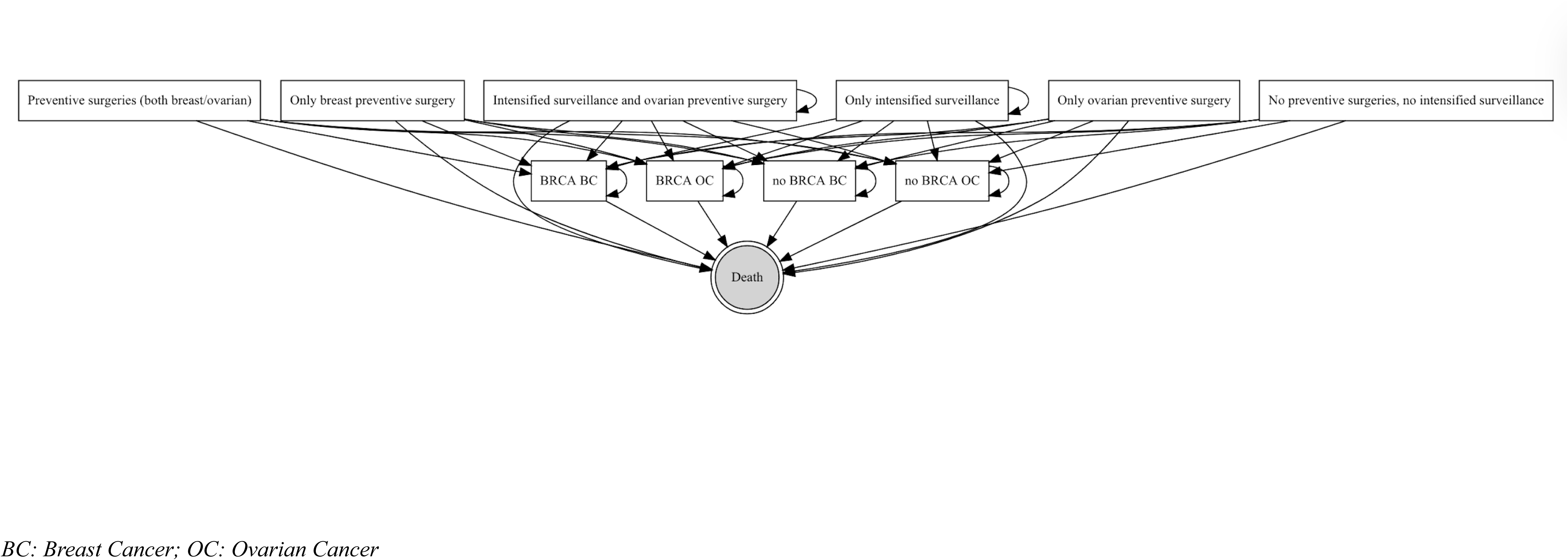
Simulation model Structure A. Markov model component.

**Table 2.**
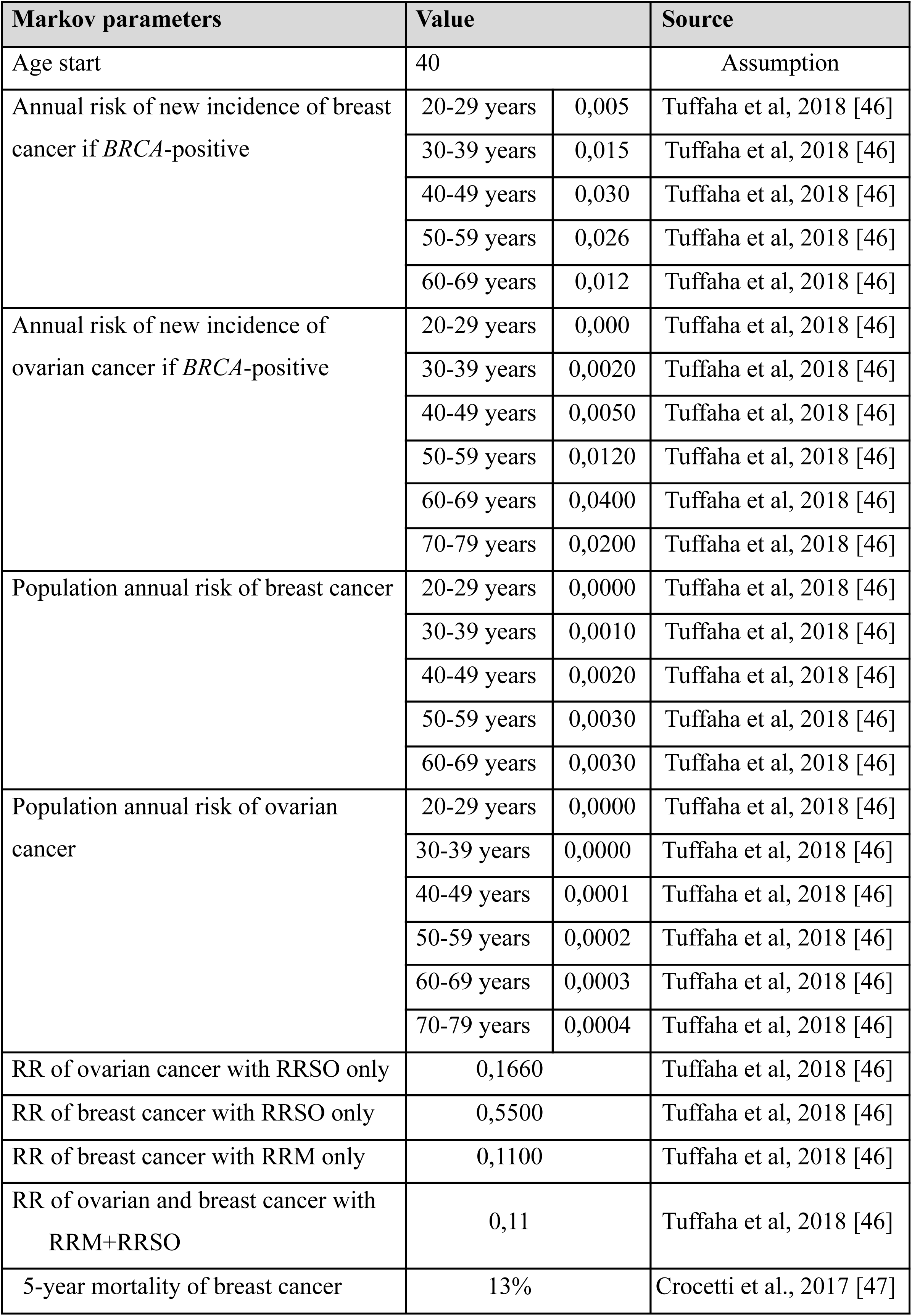
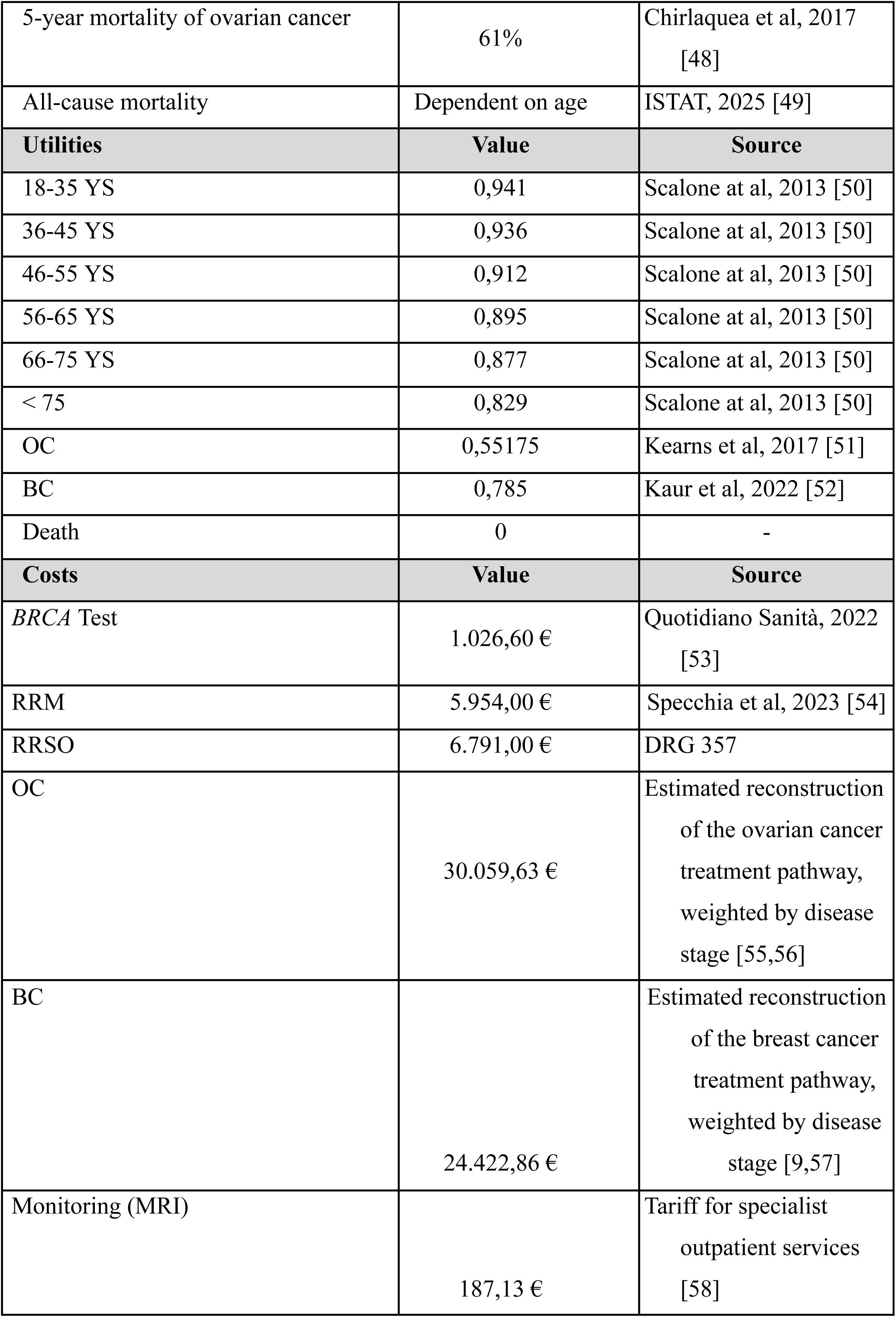
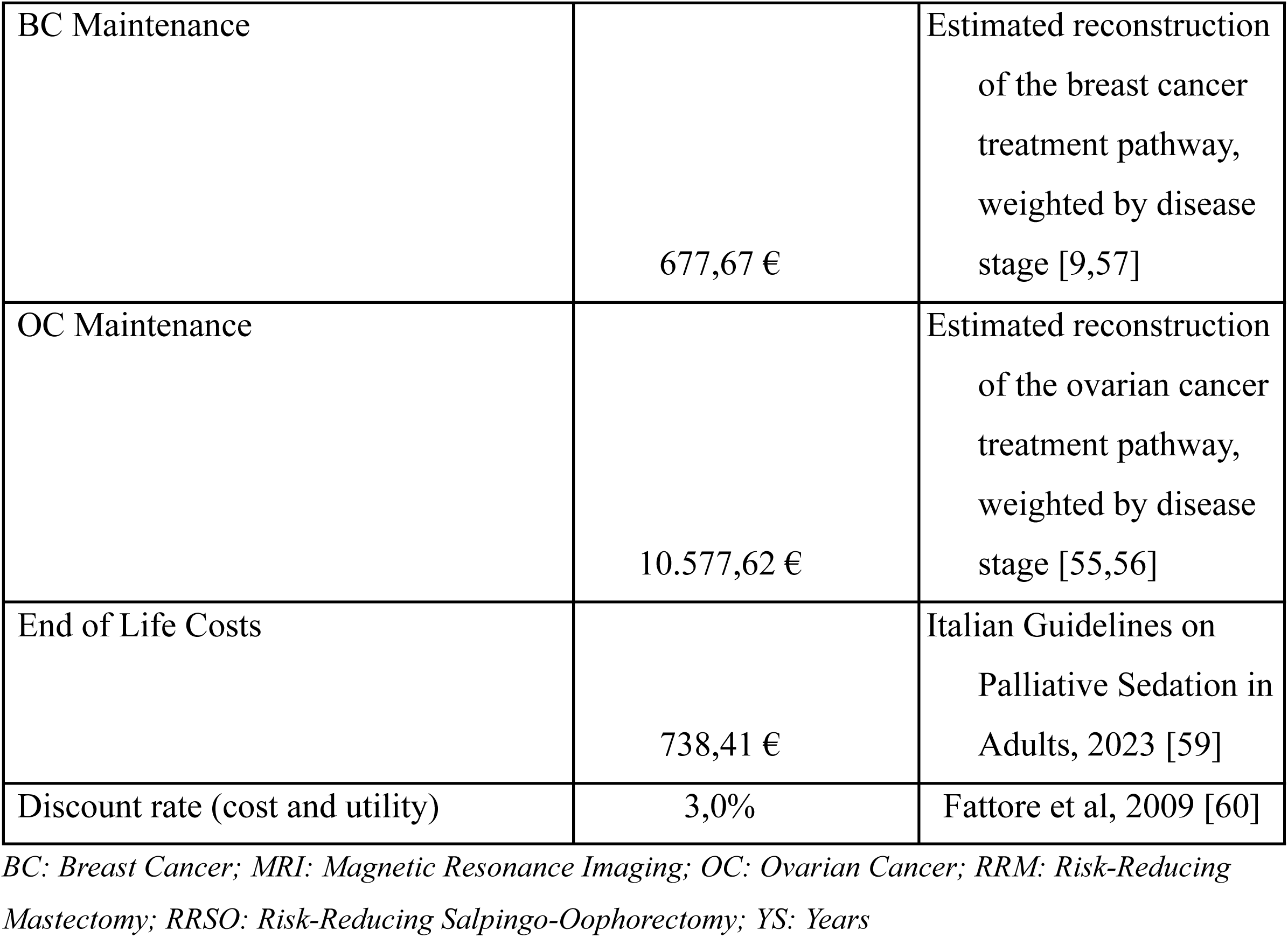
Data and sources used in the Markov model.

We estimated cumulative breast and ovarian cancer risks, as well as total costs, for each cohort at 10, 20, and 30 years. Costs for breast and ovarian cancer were estimated by calculating stage-specific diagnostic and treatment costs, in accordance with Italian guidelines, and applying them to the distribution of cases by stage at diagnosis (Supplementary Annex 1). Following the recommendations of Italian guidelines for economic evaluations, all costs were discounted at an annual rate of 3%. We also calculated the incremental cost per event avoided, representing the additional cost (or savings) associated with preventing a single case of breast or ovarian cancer or a related death.

### Recommendations

Based on the collected evidence, we developed a set of recommendations, which were subsequently discussed and validated by the SC, to improve the implementation of *BRCA* cascade screening in Italy, in alignment with the most recent national and international guidelines.

## Results

### Impact on population health

The evidence collected supports *BRCA* cascade screening as an effective preventive strategy, based on consistent findings that risk-reducing surgery markedly lowers cancer incidence and improves survival in mutation carriers [15–19]. However, the mortality benefit of active surveillance remains uncertain, despite a recent systematic review underpinning current guidelines reporting that annual MRI combined with mammography achieves a sensitivity close to 100% [17]. Modelled comparisons between an ideal implementation scenario and current practice in Italy quantify implementation gaps and their population health impact. In the ideal scenario, 25,626 eligible cancer patients would undergo *BRCA* testing annually in Italy, identifying 4,254 carriers and enabling cascade testing of 27,650 healthy relatives, with 8,682 carriers detected; assuming optimal uptake of preventive strategies, the model projected 2,127 cancer cases and 1,297 deaths at 30 years among healthy carriers (**Table 3**). In the current scenario, only 8,807 eligible patients are tested each year; cascade screening reaches 9,503 relatives, with 23% adherence, only 2,168 are tested, yielding the identification of 948 carrier relatives; under observed uptake of preventive options, this results in 2,948 cancer cases and 2,578 deaths at 30 years (**Table 3**). Overall, full implementation would avert 821 cancer cases and 1,282 deaths over 30 years (27.9% and 49.7% reductions, respectively) compared with current practice.

**Table 3.** Estimated ovarian and breast cancer cases and deaths under the ideal and the current implementation scenarios.

| **Time horizon** | **Cases Current Scenario** | **Cases Ideal Scenario** | **$\Delta$ Cases** | **Deaths Current Scenario** | **Deaths Ideal Scenario** | **$\Delta$ Deaths** |
| --- | --- | --- | --- | --- | --- | --- |
| **10 years** | 1,658 | 958 | -701 | 286 | 123 | -163 |
| **20 years** | 2,737 | 1,753 | -984 | 1,195 | 540 | -655 |
| **30 years** | 2,948 | 2,127 | -821 | 2,578 | 1,297 | -1,282 |

### Impact on Psychological Well-being and Patient Experience

The literature consistently highlights the complex psychological and experiential implications of *BRCA* genetic testing and subsequent preventive measures [20–24]. Carriers of *BRCA* mutations frequently report heightened levels of anxiety and psychological distress, particularly in the period immediately following disclosure of test results. These reactions, however, tend to be modulated by protective factors such as adequate pre-test education, effective family communication, and the opportunity to engage in an informed and deliberate decision-making process regarding preventive options [20,21]. Prophylactic surgery is associated with a reduction in perceived cancer risk and related worries, although these benefits may be accompanied by adverse effects on quality of life, particularly in relation to sexual function [22]. Psychological counselling plays a pivotal role in mitigating distress, supporting mental health, and enabling individuals to navigate the trade-offs between surveillance and surgical prevention [23]. Alongside psychological outcomes, patient satisfaction and acceptance of preventive strategies emerge as generally high, especially when care is delivered within a multidisciplinary framework that integrates oncology, genetics, surgery, and psychosocial support. Most women report positive experiences with counselling and preventive interventions, though challenges remain in ensuring that communication is tailored to individual needs. Younger women often face greater difficulty in understanding risk figures and implications, underscoring the need for age-sensitive counselling strategies. Similarly, individuals receiving inconclusive test results may struggle to interpret their cancer risk, highlighting the importance of standardised protocols to ensure clarity and consistency [24].

### Economic Impact

The literature consistently indicates that cascade screening targeting individuals at higher risk of *BRCA* mutations represents a cost-effective strategy, as it allows early intervention on hereditary cases of breast and ovarian cancer [25–29]. Based on our simulation, optimal implementation of cascade screening and subsequent preventive strategies would initially increase expenditures (due to the higher uptake of genetic testing, prophylactic interventions, and annual MRI surveillance) with an estimated additional cost of €55.5 million compared with the current scenario, but these costs would be offset in the long term by savings from reduced treatment of new breast and ovarian cancer cases. In the ideal scenario, cumulative expenditures are estimated at €97.3 million over 10 years compared with €63.3 million in the current scenario, corresponding to an incremental cost of €33.9 million **(Table 4)**. At 20 years, the additional cost decreases to €10.7 million, while at 30 years it turns into net savings of €5.8 million. At 30 years, this is reflected in a saving cost per event avoided (–€2,779), indicating that preventing cancer cases or deaths from breast or ovarian cancer results in net savings for the health system.

**Table 4.** Estimated costs under the ideal and the current implementation scenarios.

| **Time horizon** | **Cumulative cost**<br>**Ideal scenario** | **Cumulative cost**<br>**Current scenario** | **Δ Cost (€)** | **Cost (or saving) per event avoided (€)\*** |
| --- | --- | --- | --- | --- |
| **10 years** | 97.3 M | 63.3 M | +33.9 M | 39,347.50 |
| **20 years** | 128.2 M | 117.4 M | +10.7 M | 6,557.85 |
| **30 years** | 160.0 M | 165.8 M | −5.8 M | −2,779.47 |
| * $\frac{\text{Cost (€)}}{\Delta \text{Deaths (table 3)}}$ | | | | |

### Organisational feasibility

The literature identifies multiple organisational barriers that continue to hinder the effective and equitable implementation of *BRCA* testing in Italy [30–36].

The first area of concern relates to the regulatory and professional framework for genetic counselling. Although clinical genetics is formally recognised as a speciality and both clinical geneticists and oncologists are authorised to provide counselling and order germline tests, the role of genetic counsellors has not yet been formally established within the Italian health system. This lack of recognition constrains the expansion of counselling services and contributes to regional variability in access. National regulations stipulate that counselling must be provided both before and after genetic testing, ensuring that minimum standards are met; however, the uneven distribution of professionals and the limited use of telemedicine mean that in practice, access to timely and comprehensive counselling is far from uniform across the country.

A second group of challenges concerns the delivery of testing services and laboratory practices. Surveys of Italian breast and oncology centres [31] have shown marked heterogeneity in workflows, with some institutions able to provide urgent testing within tight timelines, while others report significant delays. Differences emerge in whether testing is offered pre- or post-surgery, and in the availability of pre-test counselling, leading to inconsistencies in patient pathways.

At the laboratory level, next-generation sequencing has become the standard method for *BRCA* testing, but the lack of fully standardised pipelines creates variability in results. Laboratories often analyse *BRCA* genes within broader panels, apply different bioinformatic tools, and use a variety of sequencing machines, all of which can compromise the reproducibility and comparability of results. These issues underline the need for national-level coordination and harmonisation of laboratory protocols, without which disparities in diagnostic accuracy and timeliness are likely to persist.

A third set of organisational issues is linked to the preparedness of healthcare professionals and the structure of referral pathways. Several surveys have shown that many physicians have limited knowledge of genetic testing and are not fully confident in counselling patients about its clinical utility or cost-effectiveness. This gap in professional education not only hinders appropriate test utilisation but also reinforces attrition along the testing pathway. High-risk individuals are frequently lost between referral, counselling, and test completion, reflecting weaknesses in the coordination between oncology services, primary care, and genetic units. Programs that have attempted to integrate genetic counselling into routine oncology visits have demonstrated feasibility but also revealed that referral rates remain low and that follow-up is inconsistent.

### Recommendations

Based on the findings of this assessment, the following recommendations were developed to support more consistent and equitable implementation of *BRCA* cascade screening across Italy.

1. Regularly update national guidelines and regional diagnostic–therapeutic care pathways to reflect emerging evidence, new technologies, and evolving clinical practice, through a coordinated process led by national scientific societies and multidisciplinary experts to ensure consistency across regions.
2. Strengthen implementation monitoring through standardised indicators, routine audits, and feedback loops, so that adherence to guidelines can be measured transparently, gaps can be identified early, and corrective actions can be tracked over time.
3. Consolidate and standardise care structures for hereditary cancer management by establishing dedicated multidisciplinary teams that include oncologists, geneticists, surgeons, psychologists, and case managers, with clear roles and shared protocols to improve coordination and continuity of care
4. Expand and harmonise genetic testing capacity and pathways, including adoption of next-generation sequencing where appropriate, with quality assurance standards and clear criteria for transitioning from single-gene testing to broader multigene frameworks when clinically justified.
5. Ensure timely access to preventive surgical options by expanding surgical capacity in high-demand regions, defining standardised pathways for prophylactic and reconstructive procedures, and integrating psychological assessment and follow-up as part of routine perioperative care.
6. Develop national and regional registries to support follow-up and program evaluation, providing a common infrastructure to monitor uptake, outcomes, and long-term effectiveness, and enabling benchmarking across regions.
7. Implement interoperable digital systems to link genetic, clinical, and psychosocial information, facilitating communication across services, supporting multidisciplinary decision-making, and reducing fragmentation between genetic counselling, oncology care, and preventive services.
8. Invest in structured, continuous training for healthcare professionals through accredited education programs in hereditary cancer genetics, counselling, and communication of results, with a focus on standardising competencies across settings and reducing variability in practice.
9. Expand patient-centred counselling and support across the full pathway, including pre-test education for informed decision-making, post-test counselling to interpret risk and options, and structured support for *BRCA*-positive individuals in communicating results to at-risk relatives.
10. Reduce structural inequities in access to testing and preventive care by prioritising underserved areas through targeted investments in infrastructure, dedicated funding for workforce expansion and capacity building, and mechanisms to ensure uniform availability of services regardless of region.

## Discussion

This HIA provides a comprehensive evaluation of the potential benefits, challenges, and policy implications of achieving full implementation of *BRCA1/2* cascade screening across Italy. These findings suggest that a structured, nationwide approach to genetic testing and preventive management could yield meaningful public health benefits, with the potential to avert a considerable number of hereditary breast and ovarian cancers and to reduce associated mortality. These outcomes are aligned with previous international evidence indicating that early identification of *BRCA* mutation carriers, coupled with timely risk-reducing interventions, can markedly lower the burden of hereditary cancers [35,37–39].

From a population health perspective, the modelled estimates illustrate that scaling up testing coverage to achieve full adherence to national recommendations could prevent more than a quarter of hereditary cancer cases and half of related deaths over a 30-year period. These projections confirm the strong preventive potential of *BRCA* cascade screening and its role in reducing premature mortality, particularly among women in reproductive and middle age.

The economic analysis results further reinforce the rationale for nationwide implementation. While initial costs are expected to rise due to expanded testing, counselling, and preventive interventions, these investments are projected to translate into long-term savings as cancer incidence declines. These findings are consistent with international cost-effectiveness studies demonstrating that cascade testing, particularly when integrated into public health frameworks, offers one of the most efficient approaches to hereditary cancer prevention [25–29]. The results also align with broader European evidence showing that pre-symptomatic genetic testing and prophylactic surgery generate cost savings through avoided cancer treatment and follow-up expenditures [40,41].

Beyond its quantitative findings, this HIA approach demonstrates the value of using an integrated methodological approach to evaluate complex preventive strategies such as *BRCA* cascade screening. By combining epidemiological modelling, literature synthesis, and stakeholder consultation, the HIA was able to capture not only the expected reductions in cancer burden but also the organisational, ethical, and equity dimensions that determine the real-world feasibility of implementation. This multidimensional perspective extends beyond traditional economic or clinical evaluations, providing a structured framework for understanding how preventive genomic interventions interact with existing health systems and social contexts [42–44].

The recent agreement reached by the Italian State-Regions Conference in October 2025 to include *BRCA* testing and surveillance for hereditary breast and ovarian cancer in the updated LEA represents an initial step in formalising national recognition of these preventive services [11]. Although not yet fully operational, this development signals a growing policy commitment to standardise access to *BRCA* cascade screening. In this evolving context, our findings provide evidence to support and organisational benchmarks to guide the effective and equitable translation of these provisions into practice, underscoring that structured and standardised strategies are crucial to maximising population health benefits while promoting equity.

This study has several limitations that should be acknowledged. First, the simulation model necessarily relied on secondary data sources, including estimates derived from international datasets such as SEER, due to the lack of individual-level Italian data. Although this approach ensured analytical completeness, it introduces uncertainty related to population comparability. Second, the heterogeneity of regional data on *BRCA* testing uptake and preventive surgery limited the precision of the estimates, potentially under- or over-estimating true implementation levels. Third, the literature on patient acceptability and psychological outcomes is still limited, particularly within the Italian context. Finally, while the HIA framework allows for multidimensional evaluation, some contextual factors, such as sociocultural attitudes, health literacy, and physician referral behaviour, could not be fully captured within the study.

Despite these limitations, this study represents, to our knowledge, the first comprehensive HIA conducted on the nationwide implementation of *BRCA* cascade screening in Italy. It offers an unprecedented, holistic assessment of the policy’s potential health, economic, and organisational implications. The study provides actionable evidence to inform national strategies on hereditary cancer prevention and demonstrates the utility of the HIA methodology as a framework for evaluating personalised prevention policies.

## Conclusion

Findings from this HIA suggest that the full implementation of *BRCA* cascade screening in Italy could yield substantial benefits in cancer prevention and survival, with favourable long-term economic implications. The resulting recommendations translate identified gaps into actionable priorities for policy and service delivery, offering a roadmap for more consistent implementation across the country. Beyond its national relevance, this assessment contributes to the broader European discourse on precision public health, offering a replicable model for assessing and implementing genomic screening programs aimed at equity, sustainability, and measurable population health gains.

## Availability of data and materials

The datasets used and/or analysed during the current study are available from the corresponding author on reasonable request.

## Acknowledgements

We thank the members of the Steering Committee for their valuable contributions. The Steering Committee included: Paolo Belli, Ivana Cattaneo, Alba Di Leone, Emanuela Lucci Cordisco, Aldo Rosano, Marzia Salgarello, Salvatore Testa, Alessia Tognetto.

