## Supplementary files for "Health Impact Assessment of BRCA1/2 Cascade Screening for the Personalized Prevention of Hereditary Breast and Ovarian Cancers in Italy"

**Supplementary Table 1. Criteria Applied to Establish the High-Risk Population Eligible for *BRCA* Testing.**

| **Category** | **Description** | **Proportion of cancer patients meeting each criterion*[44]** | **Source** |
|---|---|---|---|
| **Personal history** | Male breast cancer | 0,76% | SEER database [12] |
| | Woman with breast cancer and ovarian cancer | 0,60% | SEER database [11] |
| | Woman with breast cancer <40 years | 4,6% | SEER database [11] |
| | Woman with breast cancer <36 years (regional criterion)# | 2,6% | SEER database [11] |
| | Woman with triple-negative breast cancer | 8,6% | SEER database [11] |
| | Woman with bilateral breast cancer <50 years | 0,1% | SEER database [11] |
| | Woman with non-mucinous and non-borderline ovarian cancer | 97,2% | SEER database [11] |
| | Metastatic pancreatic adenocarcinoma | 49,4% | SEER database [11] |
| | Metastatic prostate cancer | 12,3% | SEER database [11] |
| **Personal history of breast cancer < 50 years** | First degree relative with Breast cancer <50 years | 0,44% | Rawal et al., 2006 [44] |
| | First degree relative with non-mucinous and non-borderline ovarian cancer | 0,1% | Rawal et al., 2006 [44] |

| | | | |
|---|---|---|---|
| | First degree relative with pancreatic cancer | 0,2% | Rawal et al., 2006 [44] |
| | First degree relative with prostatic cancer | 0,3% | Rawal et al., 2006 [44] |
| **Personal history of breast cancer > 50 years** | Family history of breast, ovarian, pancreatic or prostatic cancer in ≥2 first-degree relatives | 1,2% | Zhang et al., 2017 [45] |

*The percentage is calculated relative to the reference cancer, e.g., male breast cancer is estimated to account for 0.78% of all breast cancer cases.

**Supplementary Table 2. Eligibility Criteria for *BRCA* test on high-risk patients in the Italian region according to the Regional PDTA.**

| **Category** | **Description** | **Regions reporting the criterion in the regional PDTA*** | **N° Region** |
|---|---|---|---|
| **Personal history** | Male breast cancer | Abruzzo, Aosta Valley, Apulia, Basilicata, Calabria, Campania, Emilia-Romagna, Lazio, Liguria, Lombardy, Marche, Piedmont, Sardinia, Sicily, Tuscany, Trentino-Alto Adige, Veneto. | 17 |
| | Woman with breast cancer and ovarian cancer | Abruzzo, Aosta Valley, Apulia, Basilicata, Calabria, Campania, Emilia-Romagna, Lazio, Liguria, Lombardy, Marche, Piedmont, Sardinia, Sicily, Tuscany, Trentino-Alto Adige, Veneto. | 17 |
| | Woman with breast cancer <40 years° | None | 0 |
| | Woman with breast cancer <36 years | Abruzzo, Aosta Valley, Apulia, Basilicata, Calabria, Campania, Emilia-Romagna, Lazio, Liguria, Lombardy, Piedmont, Sardinia, Sicily, Tuscany, Trentino-Alto Adige, Veneto. | 16 |
| | Woman with breast cancer <30years | Marche | |
| | Woman with triple-negative breast cancer | Abruzzo, Aosta Valley, Apulia, Basilicata, Calabria, Campania, Emilia-Romagna, Lazio, Liguria, Lombardy, Marche, Piedmont, Sardinia, Sicily, Tuscany, Trentino-Alto Adige, Veneto. | 17 |

| | | | |
|---|---|---|---|
| | Woman with bilateral breast cancer <50 years | Abruzzo, Aosta Valley, Apulia, Basilicata, Calabria, Campania, Emilia-Romagna, Lazio, Liguria, Lombardy, Marche, Piedmont, Sardinia, Sicily, Tuscany, Trentino-Alto Adige, Veneto. | 17 |
| | Woman with non-mucinous and non-borderline ovarian cancer | Calabria, Campania, Emilia-Romagna, Lazio, Lombardia, Marche, Trentino-Alto Adige | 7 |
| | Metastatic pancreatic adenocarcinoma | Campania | 1 |
| | Metastatic prostate cancer | Campania | 1 |
| **Personal history of breast cancer < 50 years** | First degree relative with Breast cancer <50 years | Abruzzo, Aosta Valley, Apulia, Basilicata, Calabria, Campania, Emilia-Romagna, Lazio, Liguria, Lombardy, Marche, Piedmont, Sardinia, Sicily, Tuscany, Trentino-Alto Adige, Veneto. | 17 |
| | First degree relative with non-mucinous and non-borderline ovarian cancer | Abruzzo, Aosta Valley, Apulia, Basilicata, Calabria, Campania, Emilia-Romagna, Lazio, Liguria, Lombardy, Marche, Piedmont, Sardinia, Sicily, Tuscany, Trentino-Alto Adige, Veneto. | 17 |
| | First degree relative with pancreatic cancer | Calabria, Campania, Toscana | 3 |
| | First degree relative with prostatic cancer | Calabria, Campania, Toscana | 3 |

| | | | |
|---|---|---|---|
| **Personal history of breast cancer > 50 years** | Family history of breast, ovarian in ≥2 first-degree relatives | Abruzzo, Calabria, Campania, Emilia-Romagna, Liguria, Puglia, Sardegna, Toscana, Valle d’Aosta | 9 |

**Costs of ovarian cancer treatment**

We estimated the costs of ovarian cancer treatment based on stage-specific therapeutic pathways, incorporating diagnostic procedures, surgical interventions, chemotherapy, and maintenance therapy when applicable. The stage distribution was assumed to be: Stage I – 25%, Stage II – 15%, Stage III–IV – 60% [1–3]. All treatment pathways were derived from the most recent AIOM (A*ssociazione Italiana di Oncologia Medica*) clinical guidelines [4].

For Stage I patients (IA/IB G1), treatment includes non-malignant surgical procedures on the uterus and adnexa (code 359 13 C), along with diagnostic assessments such as endometrial biopsy and CA125 testing before and after surgery. In other Stage I cases, adjuvant chemotherapy with carboplatin and paclitaxel is administered every three weeks for 3–6 cycles. Follow-up includes CA125 testing and biopsy every 3 months for the first 2 years, then annually up to 6 years.

Stage II treatment includes the same surgical approach (359 13 C), followed by six cycles of adjuvant chemotherapy with carboplatin (AUC 5–6) and paclitaxel (175 $mg/m^2$) every 3 weeks. Patients undergo CA125 testing every 3–6 months for the first 2 years.

For Stage III patients, cytoreductive surgery (code 357 13 C for malignant ovarian procedures) is followed by six cycles of carboplatin and paclitaxel every 3 weeks. Maintenance therapy with bevacizumab and PARP inhibitors (in BRCA-positive cases) may be administered every 3 weeks for up to 15 months based on individual clinical indications. Follow-up includes lower abdominal ultrasound, biopsy, and CA125 testing.

Stage IV treatment consists primarily of systemic chemotherapy with carboplatin and paclitaxel, combined with maintenance therapy (bevacizumab and PARP inhibitors for BRCA-positive patients) administered every 3 weeks up to 15 months, as clinically indicated. Cytoreductive surgery may also be performed. Follow-up includes CA125 testing every 3 months, lower abdominal CT scan, and biopsy.

### Costs of breast cancer treatment

We estimated breast cancer treatment costs based on clinical stage at diagnosis and corresponding therapeutic strategies, including diagnostic procedures, surgery, radiotherapy, chemotherapy, hormonal and targeted therapy, and follow-up. The stage distribution was assumed to be: Stage 0 (in situ) – 0.6%, Stage I – 50.4%, Stage II – 32%, Stage III – 11%, and Stage IV – 6% [5]. All treatment pathways were derived from the latest AIOM (Associazione Italiana di Oncologia Medica) clinical guidelines [6].

Stage 0 (in situ) patients typically undergo breast-conserving surgery (DRG 260) and, when indicated, adjuvant radiotherapy five times per week for 5–6 weeks. Diagnostic and follow-up procedures include excisional biopsy, annual mammography, and complete blood count.

Stage I treatment includes surgery and adjuvant radiotherapy (five sessions per week for 5–6 weeks). Depending on the tumor characteristics, patients may also receive daily hormonal therapy for 5 years. Follow-up includes annual mammography, complete blood count, CA15-3 testing every 3–6 months, and clinical visits every 3–6 months for 2 years, then every 6–12 months. Sentinel lymph node biopsy is performed when clinically indicated.

Stage II patients undergo surgery followed by adjuvant chemotherapy (administered every 2–3 weeks for 4–6 cycles). Radiotherapy (5 sessions per week for 5–6 weeks) and daily hormonal therapy for 5–10 years are added based on clinical assessment. Diagnostic and follow-up procedures include biopsy, sentinel lymph node biopsy, annual mammography, complete blood count, and CA15-3 testing every 3–6 months.

Stage III treatment involves neoadjuvant chemotherapy (every 3 weeks for 4–6 months), surgery, and radiotherapy (5 sessions per week for 5–6 weeks). Hormonal therapy is prescribed daily for 5–10 years when indicated. Follow-up includes upper abdominal CT scan, bilateral breast MRI with and without contrast, CA15-3 testing, biopsy, and complete blood count.

Stage IV patients receive systemic chemotherapy every 3 weeks, targeted biological therapy every 3 weeks, and daily hormonal therapy. Monitoring includes imaging studies, biopsy, complete blood count, and CA15-3 testing every 3 months.
